# MRCAT: A multi-agent framework for reliable Mendelian randomization analysis and causal interpretation

**DOI:** 10.64898/2026.11.16.26368234

**Authors:** Yuxin Jiang, Fan Zhang, Xinrui Huang, Zhiwei Wang, Xianghong Hu, Can Yang

## Abstract

Mendelian randomization (MR) uses genetic variants to estimate causal effects of exposures on health outcomes, but rigorous MR analyses require coordinated procedures and interpretation under explicit methodological assumptions. Although artificial intelligence (AI) agents can automate analyses, computational completion does not guarantee scientific reliability. Here, we present MRCAT, a multi-agent framework that translates MR-specific knowledge into an AI-assisted workflow that is plannable, executable, verifiable, recoverable and traceable. MRCAT combines dependency-aware planning, Skill-guided execution and expertinformed review with traceable reporting guided by STROBE-MR, a guideline for clear and transparent reporting of MR studies, and iterative Writer–Judge checking. We evaluated MRCAT on 30 natural-language queries spanning diverse data sources, analytical requirements and boundary conditions. MRCAT achieved scientific completion, defined as meeting all nine prespecified MR-specific procedural requirements, for all 26 executable queries and appropriately handled all four boundary cases, compared with 17, 11 and 4 scientifically completed queries for Claude Code, Biomni and BioMedAgent, respectively. Across six independently curated tasks for which the expert-curated analyses did not support causal effects, MRCAT showed closer numerical concordance with the expert-curated analyses and produced interpretations consistent with expertcurated conclusions in all six cases, compared with two for Claude Code and four for Biomni. Additional case studies demonstrated evidence-grounded reporting and selective reuse of analytical outputs in multi-exposure and bidirectional MR investigations. MRCAT provides a framework for reducing the operational burden of MR research while supporting reproducible analysis, assumption-consistent interpretation and traceable scientific reporting.

## Introduction

Inferring causal relationships between risk factors, or exposures, and diseases is central to health management and medical intervention [1–4]. Mendelian randomization (MR) uses genetic variants as instrumental variables to estimate the causal effects of exposures on outcomes [5–7]. The increasing availability of genomewide association study (GWAS) summary statistics has substantially expanded the range of exposure–outcome relationships that can be investigated using MR [8, 9]. However, a rigorous MR investigation involves considerably more than obtaining an effect estimate [10]. Researchers must identify suitable exposure and outcome datasets, select and validate genetic instruments, preprocess and harmonize summary statistics, choose appropriate MR methods, conduct sensitivity and diagnostic analyses, and interpret and report the findings in the context of MR assumptions and limitations [11–13]. These steps are interdependent and often require substantial methodological expertise. Moreover, analyses of new exposure–outcome pairs frequently require researchers to repeat or reconsider earlier decisions. Automating and coordinating this end-to-end workflow therefore has the potential to reduce the practical burden of MR research while improving its reproducibility and transparency.

Artificial intelligence (AI) agents provide a promising approach for automating complex biomedical work-flows [14–18]. Existing systems include agents designed around predefined analytical workflows, such as MRAgent [19], general-purpose coding agents such as Claude Code [20], and biomedical coding agents such as Biomni and BioMedAgent [21, 22]. These systems demonstrate the potential of language-model-based agents to retrieve data, write and execute code, and coordinate multiple analytical tools. Nevertheless, automation alone does not ensure scientific reliability in MR. Existing agents may execute code successfully while omitting important analytical steps, selecting inappropriate datasets or methods, mishandling allele harmonization, or producing interpretations that are not supported by the available evidence [23]. A reliable MR system must therefore address not only whether an analysis can be executed, but also whether it is scientifically appropriate and whether its conclusions are justified.

In this work, we distinguish scientific reliability into two related dimensions. Procedural reliability concerns the validity of the analytical workflow, including dataset selection, instrumental-variable construction, data preprocessing, allele harmonization, method selection, diagnostic analyses and failure handling. These steps involve multiple dependencies and cannot always be resolved by generic coding or reasoning capabilities. Inferential and reporting reliability concerns whether the resulting estimates are interpreted consistently with the analytical evidence, MR assumptions and relevant methodological guidance. Without appropriate checks, an agent may generate invalid estimates, overlook important limitations, or overstate causal claims on the basis of weak or conflicting evidence. Importantly, reliable MR automation should not be understood as guaranteeing that an inferred relationship is causally true in the underlying biology [6, 24]. Rather, it should support the valid execution, evaluation and interpretation of MR analyses under their stated assumptions.

Here, we present MRCAT, a large language model (LLM)-based multi-agent framework for scientifically reliable, end-to-end MR analysis. MRCAT incorporates MR-specific knowledge into planning, execution, review and reporting rather than relying solely on the general reasoning abilities of an LLM. Its dependencyaware Planner translates research questions into workflows that preserve MR-specific prerequisites and analytical dependencies. Its Skill-guided Execution layer provides a library of 26 MR-specific Skills, combining established implementations with explicit decision rules and validation checks to constrain and monitor LLM-generated actions. MRCAT also incorporates expert-informed review and recovery policies to assess failed or incomplete steps and determine whether an analysis should be retried, revised or terminated. This approach distinguishes computational failures from scientifically informative limitations, such as insuffiicient instruments or the absence of suitable data.

To improve the reliability of causal interpretation and reporting, MRCAT includes a Report agent that integrates execution traces, computational results and relevant literature into a traceable scientific report guided by STROBE-MR recommendations [25]. An iterative writer–judge process checks for substantive omissions, inconsistencies between numerical results and interpretations, and causal claims that are not adequately supported by the analysis. MRCAT further supports follow-up investigations through persistent workspaces and recorded analytical decisions. For each new request, the Planner evaluates which previous outputs remain scientifically appropriate for reuse and which analyses must be repeated. A graphical user interface enables users to discuss research questions, revise analytical plans and inspect the resulting code, intermediate outputs and analytical artifacts.

We evaluated MRCAT across multiple levels of the MR research cycle, including execution, analytical procedures, numerical estimates and causal interpretation. First, we compared MRCAT with Claude Code, Biomni and BioMedAgent on 30 queries covering diverse data sources and analytical requirements [20, 26–29]. MRCAT achieved the highest rates of both execution completion, defined as the generation of an MR estimate, and scientific completion, defined as fulfilment of all prespecified MR-specific evaluation criteria. It also correctly identified the predefined cases in which scientific or data limitations prevented a complete analysis. We next compared the agents with expert-curated analyses at the levels of analytical procedures, numerical estimates and causal interpretations. In a method-specific case study, MRCAT closely reproduced the expert-curated procedures and numerical estimates. Across six independently curated tasks for which the expert-curated analyses did not support causal effects, MRCAT produced consistent interpretations in all six cases, compared with two for Claude Code and four for Biomni. We further assessed MRCAT’s ability to generate evidence-grounded reports and to support scientifically justified reuse of prior analytical outputs in follow-up investigations, including analyses of candidate risk factors for rheumatoid arthritis and bidirectional relationships between type 2 diabetes and coronary artery disease.

## Results

### MRCAT embeds domain knowledge to support procedural and inferential reliability in Mendelian randomization

MRCAT is a multi-agent framework for reliable, end-to-end Mendelian randomization (MR) analysis (Fig. 1a). Users can initiate an investigation with a natural-language question or local GWAS data, and refine the request conversationally. A central Manager interprets the research objective and coordinates three specialized agents: Search, MR and Report. Search retrieves relevant literature and data to clarify the scope of the investigation. MR plans and executes the analytical workflow, including data preparation, instrument construction, estimation and diagnostics. Report integrates the executed analyses and supporting evidence into a traceable scientific report. This architecture connects research-question formulation, data acquisition, MR analysis and reporting while keeping the underlying code, results and analytical artifacts available for inspection (Fig. 1b). MRCAT supports procedural reliability through three coordinated components. First, dependency-aware planning translates the research objective and available inputs into an ordered workflow, accounting for study design, data availability and method-specific prerequisites. Second, Skill-guided execution carries out the approved workflow using 26 MR-specific Skills (Supplementary Table 1). Each Skill combines established implementations with explicit decision rules, quality checks and stopping conditions. Programmatic checks address directly testable requirements, whereas written guidance supports context-dependent methodological judgements. Custom code can be used when an operation is not fully covered by existing tools while remaining within the same execution and validation framework. Third, expert-informed review evaluates failed or blocked Skill executions, intermediate outputs and completed custom analyses against the available data and MR-specific requirements. It determines whether to correct the current step, revise upstream preparation, retry the analysis or stop at a scientifically justified boundary. This process distinguishes computational failures from informative scientific limitations, such as insuffiicient instruments, incompatible datasets or the absence of suitable data, thereby extending methodological scrutiny beyond code execution to decisions about whether and how an analysis should proceed.

**Figure 1.**
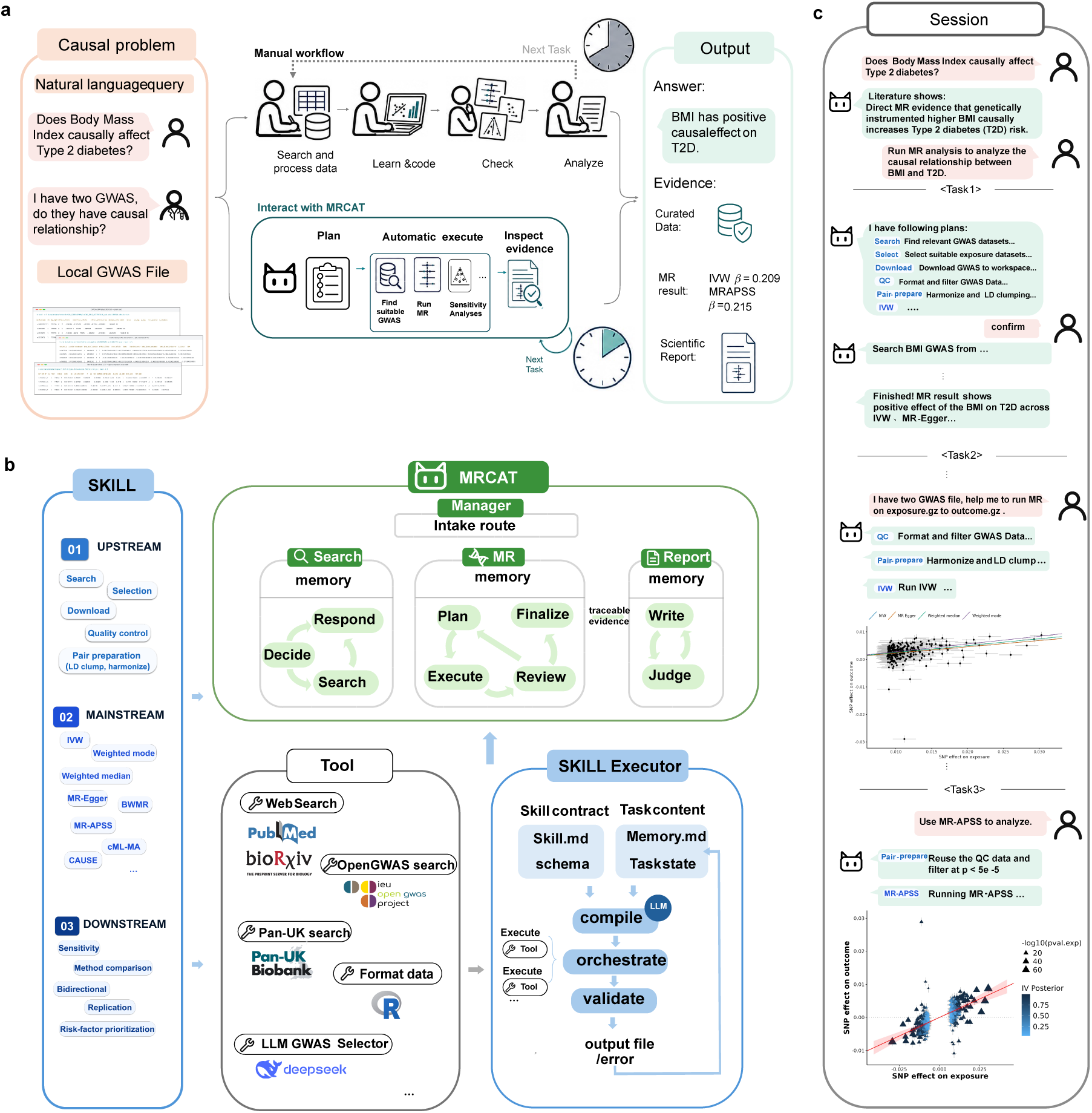
| Overview of MRCAT. a, Research workflow from natural-language questions or local genome-wide association study (GWAS) files to Mendelian randomization (MR) results and scientific reports. Manual analysis is contrasted with interactive planning, automated execution and evidence inspection using MRCAT. **b,** System architecture. The Manager coordinates Search, MR and Report workflows, including the MR Plan–Execute–Review loop and the Report Writer–Judge loop. Skills cover data preparation, MR estimation and downstream analyses; the Skill Executor combines Skill instructions with task context to execute tools and validate outputs. **c,** Illustrative session progressing from literature inquiry and online-data analysis to local-file analysis and a follow-up MR-APSS request. Quality-controlled data are reused with method-specific preparation. Body mass index (BMI) and type 2 diabetes (T2D) are example traits.

MRCAT supports inferential and reporting reliability through three corresponding functions. First, traceable evidence retrieval identifies the analyses relevant to the user’s request and assembles their estimates, diagnostic results, visualizations and provenance. Whereas Search retrieves external literature and data, this reporting layer retrieves evidence generated by the executed analysis, grounding interpretation in the actual analytical record. Second, the Writer integrates the selected evidence using MR-specific guidance and STROBE-MR recommendations [25]. Third, the Judge reviews the report for substantive omissions, inconsistencies between numerical results and their interpretation, and causal claims that are not adequately supported by the analysis or its assumptions. Targeted revision and renewed review help align the final report with the strength and limitations of the evidence. This process supports assumption-consistent interpretation rather than guaranteeing that an inferred relationship reflects a true underlying biological causal effect.

A persistent workspace preserves research context, analytical decisions and generated outputs, allowing follow-up requests to build on an ongoing investigation. In the BMI–T2D example (Fig. 1c), Search first retrieves relevant literature and data to help define the scope of the analysis. The Manager then routes the request to MR, where the Planner proposes an ordered sequence of Skill-based operations for user confirmation. After confirmation, the Executor performs the workflow, while code, estimates, diagnostic results and visualizations remain available for inspection. When local GWAS files are provided, external data retrieval is no longer an upstream prerequisite, allowing the workflow to begin with quality control and data preparation. For follow-up requests, the Planner evaluates which previous outputs remain suitable and which operations must be repeated. For example, an MR-APSS analysis [30] may reuse previously quality-controlled study data when they remain appropriate, while retaining the method-specific preparation required for estimation. This selective reuse reduces redundant preparation without assuming that prior outputs are automatically valid for a new method or research question, thereby preserving continuity while maintaining the analytical prerequisites and methodological checks required for each subsequent analysis.

### Benchmarking shows that MRCAT achieves higher procedural reliability than related AI agents across diverse MR tasks

To evaluate MRCAT’s procedural reliability relative to other AI agents, we constructed a benchmark of 30 natural-language queries comprising 26 executable queries and four boundary queries (Fig. 2a; Supplementary Table 2). The executable queries specified MR analyses for which suffiicient data and information were available. They tested whether an agent could complete the required procedures and produce an MR estimate. The boundary queries contained unavailable data or missing analytical prerequisites and therefore tested whether an agent could recognize the limitation and respond appropriately. For example, when no downloadable GWAS for trait left-handedness was available from the specified resource, an appropriate response was to report the limitation and identify the information or resource required to proceed.

**Figure 2.**
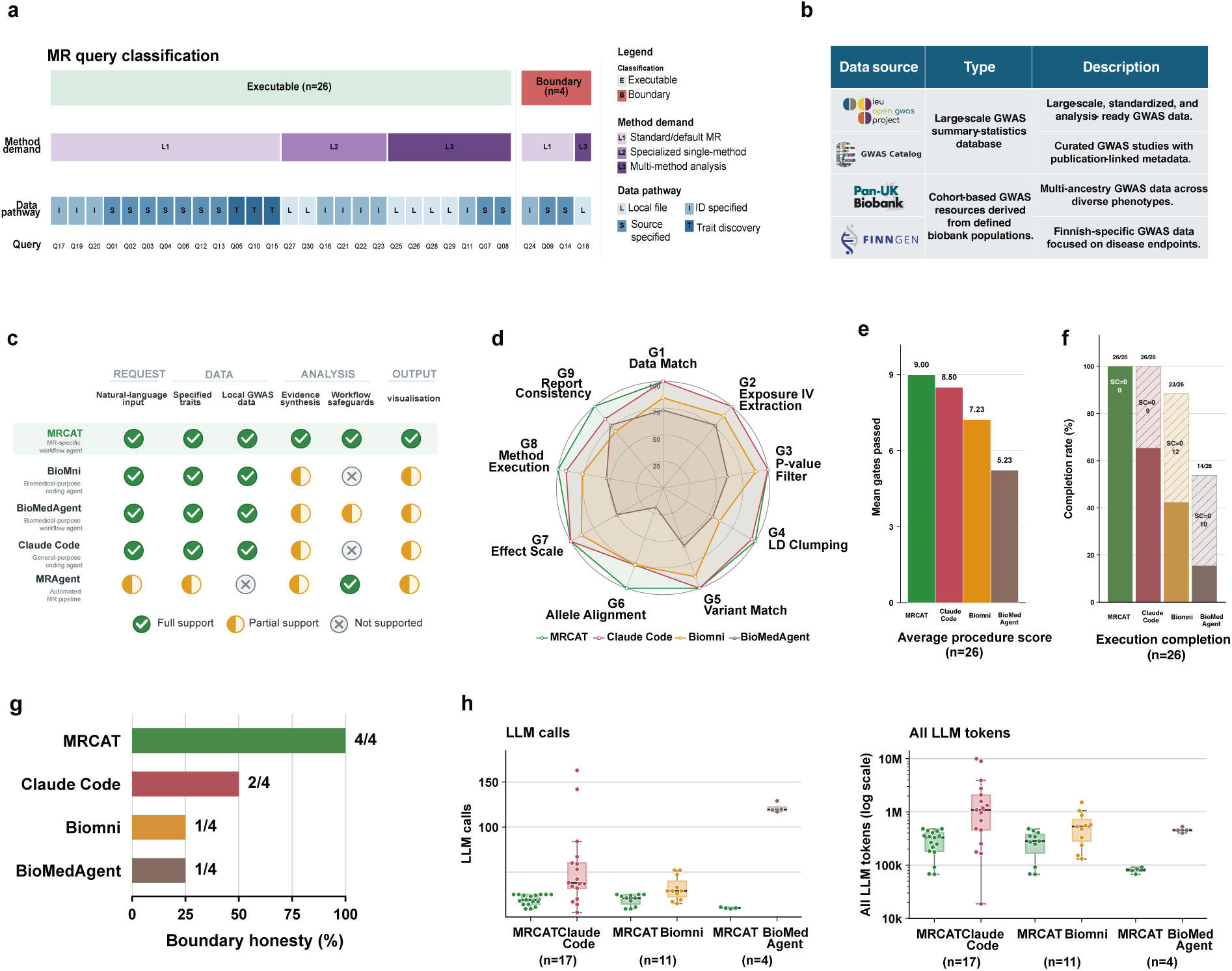
| Task coverage, completion and resource use across MR agents. a, Benchmark query design. Thirty queries comprise 26 executable tasks and four boundary cases. Method demand: L1, standard/default MR; L2, a specified specialized method; L3, multiple methods. Data pathway: L, local exposure and outcome files; I, specified study or phenotype identifiers; S, specified sources; T, trait-based discovery. **b,** Four GWAS resources represented in the benchmark: OpenGWAS, GWAS Catalog, Pan-UK Biobank and FinnGen. **c,** Capability comparison across five agents and six workflow functions. Symbols indicate full, partial or absent support. **d,** Per-gate pass rates across nine procedural requirements (G1–G9). Colours identify MRCAT (green), Claude Code (red), Biomni (orange) and BioMedAgent (brown). **e,** Mean procedure scores are 9.00, 8.50, 7.23 and 5.23. **f,** Execution completion and scientific completion among 26 executable queries. Completed-task counts are 26, 17, 11 and 4 out of 26. Solid segments indicate scientific completion; hatched segments indicate queries yielding MR estimates without scientific completion (SC=0). Labels show query counts. **g,** Boundary honesty among four boundary queries, requiring either a justified stop or a disclosed, scientifically appropriate adaptation. Labels show the number of queries meeting this criterion. **h,** Box plots of LLM calls and total tokens for matched queries where both systems passed all nine gates and had complete usage records. Comparisons with Claude Code, Biomni and BioMedAgent included 17, 11 and 4 matched pairs, respectively. The token axis is logarithmic.

The benchmark covered diverse MR scenarios involving different method requirements and data pathways. Queries either left the MR method unspecified (L1), required a single prespecified method (L2), or requested comparisons across multiple methods (L3). Inputs consisted of trait names alone (T), specified GWAS resources (S), study or phenotype identifiers (I), local files (L), or combinations of these input types. Online data requests covered OpenGWAS, the GWAS Catalog, Pan-UK Biobank and FinnGen (Fig. 2b). The complete queries and their classifications are provided in Supplementary Table 2.

We considered four existing AI agents with capabilities relevant to MR analysis: Claude Code, Biomni, BioMedAgent and MRAgent (Fig. 2c). MRAgent did not support local GWAS files, specified study identifiers or several of the requested data-source constraints and was therefore excluded from the primary fullbenchmark comparison. Its results for the six compatible queries are reported separately in Supplementary Table 3. The primary comparison included MRCAT, Claude Code, Biomni and BioMedAgent, which could process the natural-language requests and GWAS input types represented in the benchmark. These systems were evaluated using the same queries, input data, LLM backbone and underlying software packages, described in Supplementary Table 5.

Procedural compliance was assessed using nine prespecified requirements (G1–G9), covering data matching, exposure-instrument extraction, *P*-value filtering, LD clumping, variant matching, allele alignment, effect-scale handling, method execution and reporting consistency (Supplementary Table 6). An independent LLM judge, Codex with GPT-6.1 Sol, assessed the agents’ execution traces, intermediate result files and final reports according to the prespecified rubric (Methods). Judge assessments were checked against the underlying execution evidence, and the complete query-level scores are provided in Supplementary Fig. 1.

MRCAT achieved higher pass rates across the procedural gates than the comparator agents (Fig. 2d). Relative to Biomni and Claude Code, respectively, MRCAT showed pass-rate gains of 62.5% and 4.0% for LD clumping (G4), 30.0% for allele alignment (G6) and 44.4% and 18.2% for reporting consistency (G9). Inspection of execution traces and judge rationales identified recurring sources of failure. In Q26, which required multiple MR methods to be applied to local GWAS files, Claude Code replaced LD clumping with physical-distance pruning before applying cML-MA and IVW. MRCAT instead used its dependency-aware Planner to place the required harmonization and LD-clumping steps before estimation and then executed both methods using the prepared inputs. In Q30, which required MR-APSS, Biomni reversed exposure-allele labels without correspondingly updating the effect signs. MRCAT’s harmonization Skill instead preserved the correspondence between allele orientation and effect direction.

Across the 26 executable queries, MRCAT achieved a mean procedural score of 9.00 out of 9, compared with 8.50 for Claude Code, 7.23 for Biomni and 5.23 for BioMedAgent (Fig. 2e). We next distinguished execution completion, defined as producing a numerical MR effect estimate during execution, from scientific completion, defined as passing all nine procedural requirements. MRCAT and Claude Code generated estimates for all 26 executable queries, compared with 23 for Biomni and 14 for BioMedAgent. Under the stricter scientific-completion criterion, MRCAT completed all 26 queries, compared with 17 for Claude Code, 11 for Biomni and four for BioMedAgent (Fig. 2f). Among queries that generated an estimate, 9/26 (34.6%) for Claude Code, 12/23 (52.2%) for Biomni and 10/14 (71.4%) for BioMedAgent failed at least one procedural requirement. In contrast, all 26 MRCAT queries that generated estimates passed all nine requirements. Thus, generating an MR estimate did not reliably indicate completion of the required MR procedures, whereas MRCAT combined estimate generation with full procedural compliance across the executable benchmark.

We next assessed whether agents appropriately recognized and handled analytical limitations. Boundary honesty required an agent either to stop and explain the information needed to proceed or to implement a scientifically appropriate adaptation and disclose its consequences and scope (Supplementary Table 6). MR-CAT satisfied this criterion in all four boundary cases, compared with two for Claude Code and one each for Biomni and BioMedAgent (Fig. 2g). In Q18, the exposure data lacked the sample-size information required for MR-APSS. Claude Code proceeded using unsupported approximations, whereas Biomni omitted required preparation steps and generated unsupported MR-APSS inputs. BioMedAgent failed during samplesize preparation. MRCAT’s expert-informed Review instead determined that the prerequisite could not be resolved through further execution, paused the analysis and requested the information to continue. These results show that MRCAT’s review mechanism can distinguish a correctable execution problem from an unresolved scientific prerequisite and prevent unsupported analyses from proceeding.

Finally, we assessed whether MRCAT’s procedural safeguards were associated with increased computational resource use. We included only matched executable queries for which both systems passed all nine procedural gates and provided complete usage records, yielding 17 matched pairs with Claude Code, 11 with Biomni and four with BioMedAgent. MRCAT used fewer median LLM calls and tokens in each pairwise comparison (Fig. 2h). These comparisons were conditional on scientifically completed runs and therefore did not include the cumulative usage of earlier unsuccessful or incomplete runs. Together, these results show that MRCAT’s advantage was not limited to generating numerical estimates. Across diverse MR tasks, it more consistently satisfied prespecified procedural requirements, recognized scientifically meaningful limitations and maintained analytical validity through its dependency-aware planning, Skill-guided execution and expert-informed review framework.

### MRCAT closely reproduces expert-curated MR analyses and evidence-supported conclusions

We next evaluated whether the procedural reliability observed in the benchmark translated into reproducible analytical procedures, numerical concordance and reliable causal conclusions. We used two complementary sets of expert-curated MR analyses. First, we selected the BMI–type 2 diabetes (T2D) example from the MR-APSS tutorial to assess whether agents could reproduce a specified analytical workflow and its numerical results. Second, we selected six validated negative-control exposure–outcome pairs from MRbenchmarking [31]. These pairs were accompanied by GWAS data and detailed, method-specific results and therefore enabled us to assess both numerical agreement and consistency with the expected absence of a causal effect. Both resources provided detailed, publicly accessible expert-curated analyses, making them suitable standards for evaluating agent performance on these tasks.

We first assessed whether the agents reproduced the expert-curated workflow for the BMI–T2D MR-APSS analysis. Agents were provided with local GWAS summary statistics and asked to perform the analysis using MR-APSS (Supplementary Table 9). We compared their execution traces with the expert-curated workflow, including data formatting, background-parameter estimation, allele harmonization, LD clumping and causal-effect estimation (Fig. 3a). This stepwise comparison allowed us to identify procedural deviations and examine how they affected the resulting numerical estimates.

**Figure 3.**
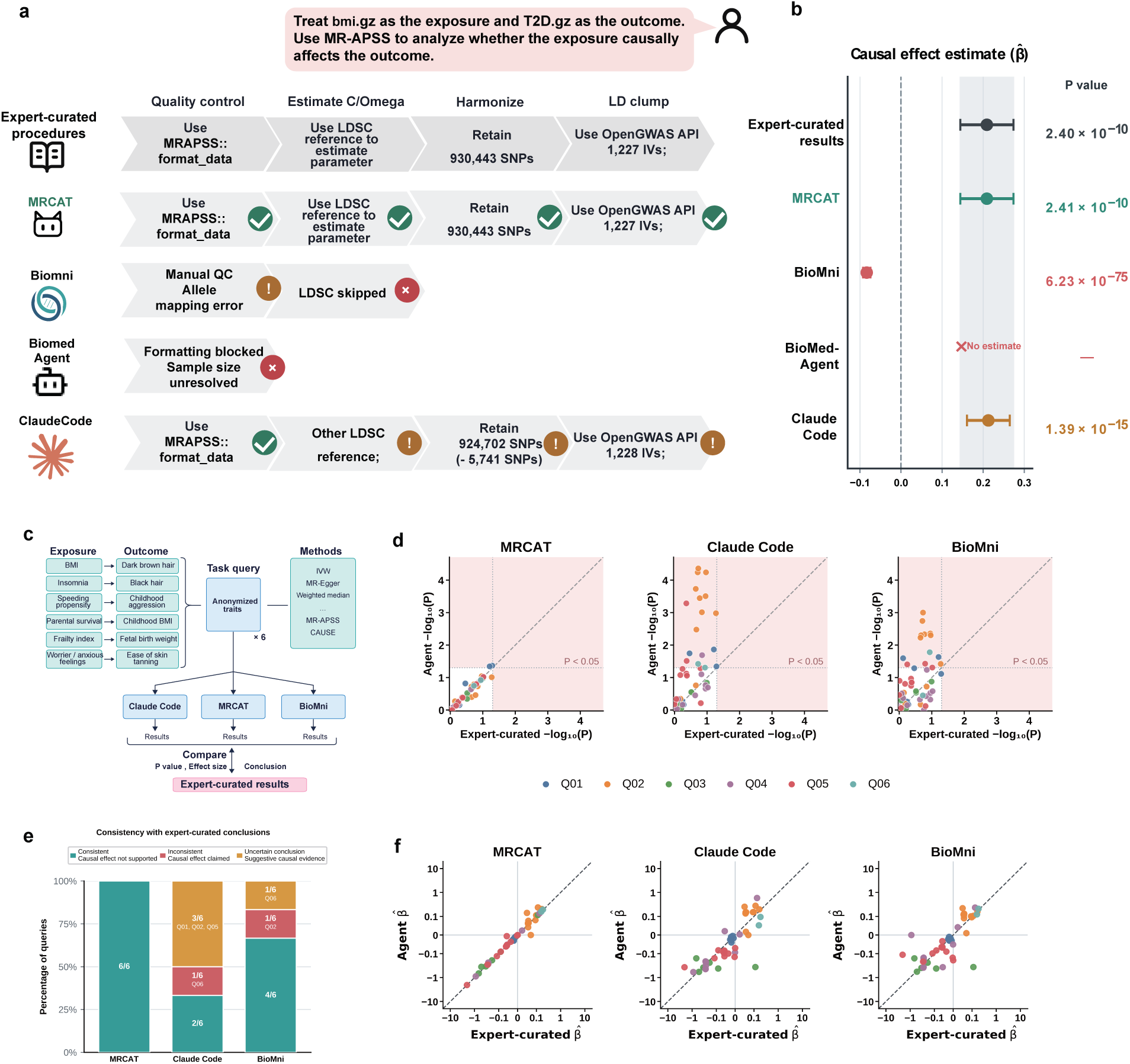
| Concordance of analytical execution and causal interpretation with expert-curated analyses. a, Stepwise comparison of agent workflows with an expert-curated MR-APSS analysis of local BMI and T2D summary statistics. Steps cover data formatting, background-parameter estimation, harmonization and LD clumping. **b,** Corresponding MR-APSS effect estimates and reported *P* values. Points and horizontal bars show estimates and 95% confidence intervals; the shaded band shows the expert-curated confidence interval. BioMedAgent produced no estimate. **c,** Evaluation of six negative-control exposure–outcome pairs with anonymized trait labels. Agent outputs were compared with expert-curated results from MRbenchmarking [31]. **d,** Agent versus expert-curated *−* log_10_(*P*) values. Dotted lines mark *P* = 0.05. Pink shading indicates *P <* 0.05 for the agent (above the horizontal line) or expert-curated results (right of the vertical line). **e,** Final conclusions across six queries per system, classified as consistent with expert-curated conclusions (causal effect not supported), inconsistent with expert-curated conclusions (causal effect claimed) or uncertain (suggestive causal evidence). Counts in the consistent category are 6*/*6, 2*/*6 and 4*/*6 for MRCAT, Claude Code and Biomni, respectively. **f,** Agent versus expert-curated effect estimates on symmetric logarithmic axes. In **d,f**, each point represents one query–method result, colours identify queries and dashed diagonals indicate equality.

MRCAT reproduced the expert-curated workflow, retaining 1,227 instruments and obtaining an effect estimate of β^^^ = 0.209, with SE = 0.033 and *P* = 2.41 *×* 10*^−^*^10^, consistent with the expert-curated results (Fig. 3b). The comparator agents deviated from the expert-curated procedure in ways that affected their numerical outputs. Claude Code used a different LD-score reference and retained 1,228 instruments. Although its effect estimate was similar to the expert-curated results (β^^^ = 0.213), it produced a smaller standard error of 0.0266 and a substantially different *P* value (1.39 *×* 10*^−^*^15^). Biomni incorrectly mapped exposure-effect alleles, replaced background-parameter estimation with fixed values and omitted LD clumping. It consequently fitted 7,442 variants selected solely by association significance and obtained an estimate in the opposite direction (β^^^ = *−*0.084, *P* = 6.23 *×* 10*^−^*^75^), which its final report interpreted as strong evidence of a negative causal effect. BioMedAgent failed to resolve the exposure sample-size field and did not produce a valid estimate. These comparisons illustrate how deviations in upstream analytical procedures can alter numerical estimates, whereas MRCAT maintained agreement with the expert-curated workflow and numerical results.

Reproducing an analysis that provides evidence compatible with a causal effect is only one aspect of reliability. Additional positive-control examples are presented in Supplementary Table 10. Inferential reliability also requires agents to avoid causal claims when the evidence does not support them. To assess this capability, we used six validated negative-control exposure-outcome pairs from MRbenchmarking (Fig. 3c). For each task, agents were provided with local GWAS summary statistics and asked to apply a prespecified set of MR methods that did not support a causal effect in the corresponding expert-curated analyses. Trait labels were anonymized in the queries to reduce reliance on phenotype-specific prior knowledge during analysis and interpretation. We extracted the method-specific effect estimates and *P* values reported in each final solution and recorded the agent’s overall causal conclusions. BioMedAgent did not provide complete final reports containing the requested numerical results for the six tasks (Supplementary Table 14).

MRCAT produced numerical results that were closer to the corresponding expert-curated analyses than those produced by the comparator agents, including for method-specific effect estimates and *P* values (Fig. 3d,f; Supplementary Table 11). Among the 53 prespecified non-significant method-specific expert-curated results, only two MRCAT results reached nominal significance, compared with 17 for Claude Code and 14 for Biomni. For example, in Q02, MRCAT’s IVW estimate closely matched the expert-curated results in both effect size and statistical significance (β^^^ = 0.063, *P* = 0.169 versus β^^^ = 0.057, *P* = 0.194). Its transparent execution records, accessible through its interface, showed that its stricter MAF filter during quality control (*MAF ≥* 0.05 versus *≥* 0.01 in the expert-curated analysis) contributed to this small difference. Claude Code instead returned larger effect estimates (β^^^ = 0.146) with much smaller *P* values (5.76 *×* 10*^−^*^5^; Supplementary Table 11). In this analysis, Claude Code converted a non-significant expert-curated result into apparent false-positive statistical evidence. Examination of the analysis traces showed that Claude Code had incorrectly reversed the outcome-effect signs for six palindromic SNPs during harmonization. Correcting these six signs changed all nine previously significant method-specific results to non-significant (Supplementary Table 13). By contrast, the two nominally significant MRCAT results were close to their corresponding expert-curated values: *P* = 0.043 versus 0.051 for the weighted median and *P* = 0.045 versus 0.062 for MR-ConMix (Supplementary Table 11). More stringent MAF filtering and, for the weighted median, bootstrap variability also contributed to these small numerical differences. More importantly, MRCAT did not interpret these isolated nominally significant results as suffiicient evidence of a causal effect, demonstrating inferential reliability in these tasks (Supplementary Table 12).

MRCAT’s final conclusions were consistent with the negative-control expectation that the analyses should not support a causal effect in all six tasks, compared with two tasks for Claude Code and four for Biomni (Fig. 3e; Supplementary Table 12). Claude Code stated in Q06 that “Both methods indicate a positive causal effect of the exposure on the outcome”, whereas Biomni concluded in Q02 that “the overall evidence favors a positive causal effect”, citing agreement across multiple methods (Supplementary Table 12). The remaining three Claude Code conclusions and one Biomni conclusion were classified as weak or uncertain. In summary, MRCAT showed closer numerical concordance with the expert-curated analyses and greater consistency with their evidence-supported interpretations (Fig. 3d–f; Supplementary Tables 11 and 12).

### MRCAT integrates analytical evidence into traceable and reliable scientific reports

To illustrate how MRCAT integrates analytical evidence into a traceable and scientifically reliable report, we used an interactive investigation of hexadecanedioate and coronary artery disease (CAD). The user first requested a two-sample MR analysis, then requested a replication analysis using FinnGen as an alternative outcome cohort, and finally asked MRCAT to integrate and interpret the accumulated findings. Although the effect estimates were consistently negative, the statistical support for a protective effect varied across MR methods and outcome cohorts. The report therefore needed to preserve these differences and distinguish consistent effect directions from reproducible evidence for causality. MRCAT generated the report through its Writer–Judge workflow, guided by MR-specific interpretation rules and STROBE-MR recommendations (Fig. 4a,b). The complete report is provided in the Supplementary Methods and Results under “Complete scientific report”.

**Figure 4.**
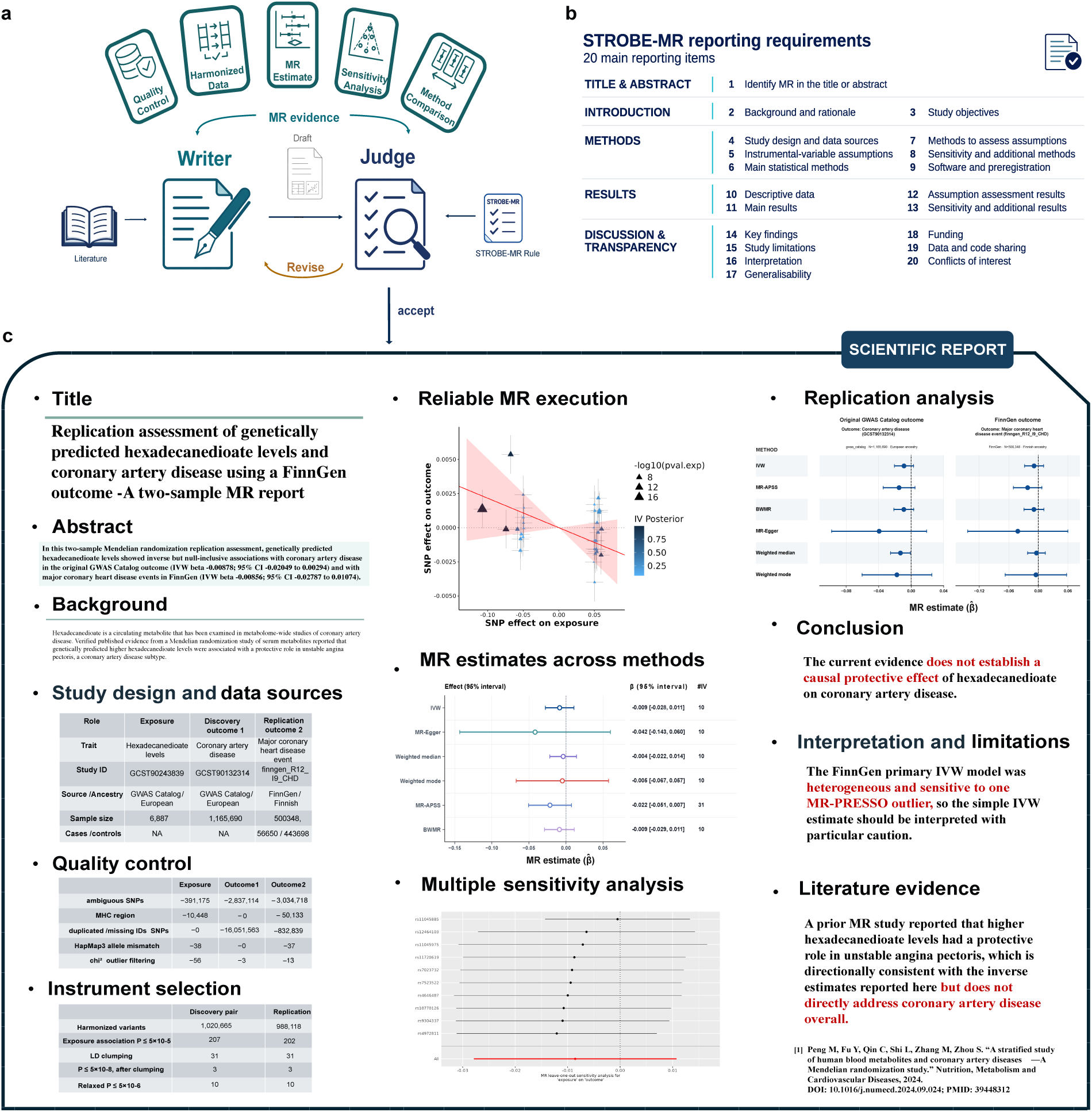
| Evidence integration and review for MR report generation. a, Writer–Judge workflow. The Writer integrates analytical evidence and literature into a draft. The Judge checks reporting completeness against STROBE-MR guidance, result–interpretation consistency and support for causal claims, returning the draft for revision or accepting it for delivery. **b,** The 20 main STROBE-MR reporting items provided to support the reporting of MR studies. **c,** Selected report content for genetically predicted circulating hexadecanedioate and coronary artery disease, including a FinnGen analysis of major coronary heart disease events. The report presents data provenance, quality control, instrument selection, MR estimates, sensitivity analyses, comparison between cohorts, limitations and relevant literature. The panel illustrates report content rather than a quantitative assessment of reporting accuracy.

To make the reported findings traceable to the underlying analyses, MRCAT documented the data sources, processing decisions and results for both analysis branches. The report identified the shared exposure dataset and the two outcome datasets, including their study sources, ancestry and sample sizes (Supplementary Table 15). It summarized quality control, allele harmonization, LD clumping and instrument selection and recorded the number of variants retained at each stage (Supplementary Tables 15 and 16). These records allowed readers to follow how the input datasets were transformed into the instruments used for each analysis. Tables and figures presented effect estimates, confidence intervals, *P* values and instrument counts for six MR methods across the original CAD and FinnGen analyses, clearly distinguishing the two outcome cohorts (Fig. 4c; Supplementary Table 17). Method-comparison and replication plots displayed the results side by side, whereas leave-one-out plots showed the influence of individual instruments. By retaining both nominally significant and non-significant results, the report made the evidence supporting and limiting the causal interpretation available for inspection.

MRCAT also documented the methodological decisions and diagnostic findings relevant to interpretation. In both analysis branches, only three instruments met the stringent exposure-association threshold of *P ≤* 5 *×* 10*^−^*^8^. The threshold was therefore relaxed to *P ≤* 5 *×* 10*^−^*^6^ to retain ten instruments for the primary analysis, and the report explicitly characterized the resulting analyses as exploratory (Supplementary Table 16). It further described differences in diagnostic evidence between the two branches. The FinnGen analysis showed heterogeneity according to Cochran’s *Q* test (*P* = 0.0208) and greater sensitivity to an influential instrument, although leave-one-out analyses did not reverse the direction of the IVW estimate (Supplementary Table 16). These findings were reported alongside the consistent negative effect directions rather than being omitted from the summary. The report also presented instrument-strength measures and explicitly noted that the instrumental-variable assumptions could not be fully verified from summary data.

The report then calibrated its causal claims to the combined evidence. The confidence intervals for the primary IVW estimates included the null in both outcome datasets. Although the weighted-median estimate was nominally significant in the original analysis (*P* = 0.0344), this finding was not replicated in FinnGen (*P* = 0.6833; Supplementary Table 17). MRCAT therefore distinguished consistency in effect direction from consistent statistical evidence for a protective causal effect. The report also noted that the previous study examined unstable angina, whereas the present analyses considered broader CAD outcomes and major coronary heart disease events, limiting direct comparability with the published finding (Fig. 4c; Supplementary Methods and Results). Considering the null-inclusive confidence intervals for the estimates, the lack of replication of the nominally significant weighted-median finding, and the heterogeneity and outlier sensitivity in the FinnGen analysis, the report concluded that the available evidence was insuffiicient to establish a protective causal effect.

This case illustrates how MRCAT supports inferential and reporting reliability by linking conclusions to the underlying data, processing decisions, estimates and diagnostics. Rather than treating consistent effect directions or isolated nominally significant results as suffiicient evidence of causality, MRCAT preserves uncertainty and methodological limitations in the final report and calibrates its interpretation to the strength and reproducibility of the evidence.

### MRCAT supports downstream applications in risk-factor assessment and bidirectional analysis

MR investigations often extend beyond a single exposure–outcome analysis. Researchers may evaluate multiple candidate risk factors for the same disease or examine the reverse direction of an association. Such follow-up analyses require more than repeated execution: the system must identify data and analytical artifacts that remain suitable for reuse while preserving the dependencies specific to each exposure–outcome pair or causal direction. We illustrate these capabilities through a multi-exposure assessment of potential causal risk factors for rheumatoid arthritis (RA) and a bidirectional MR investigation of type 2 diabetes (T2D) and coronary heart disease (CHD). The selected GWAS datasets and analysis settings are listed in Supplementary Tables 18 and 19.

We first asked MRCAT to evaluate smoking, hypothyroidism, Graves’ disease, educational attainment and walking pace as potential causal risk factors for RA. MRCAT recognized the request as an MR investigation and organized it through its Planner–Executor–Review workflow. The Planner decomposed the request into a 50-step workflow covering GWAS discovery and selection, data retrieval and quality control, exposurespecific instrument selection, pair-specific harmonization, estimation using six MR methods—IVW [32], MR-Egger [33], weighted median [34], weighted mode [35], MR-APSS [30] and BWMR [2]—sensitivity analyses and cross-exposure synthesis. Rather than running five fully independent pipelines (Fig. 5a), MR-CAT reused the quality-controlled RA outcome dataset while independently selecting instruments and performing harmonization for each exposure (Fig. 5b). The Executor passed the required outputs between successive Skills, while preserving the pair-specific inputs, decisions and processing history for each analysis. Review assessed intermediate outputs and resolved or terminated unsupported steps when necessary.

**Figure 5.**
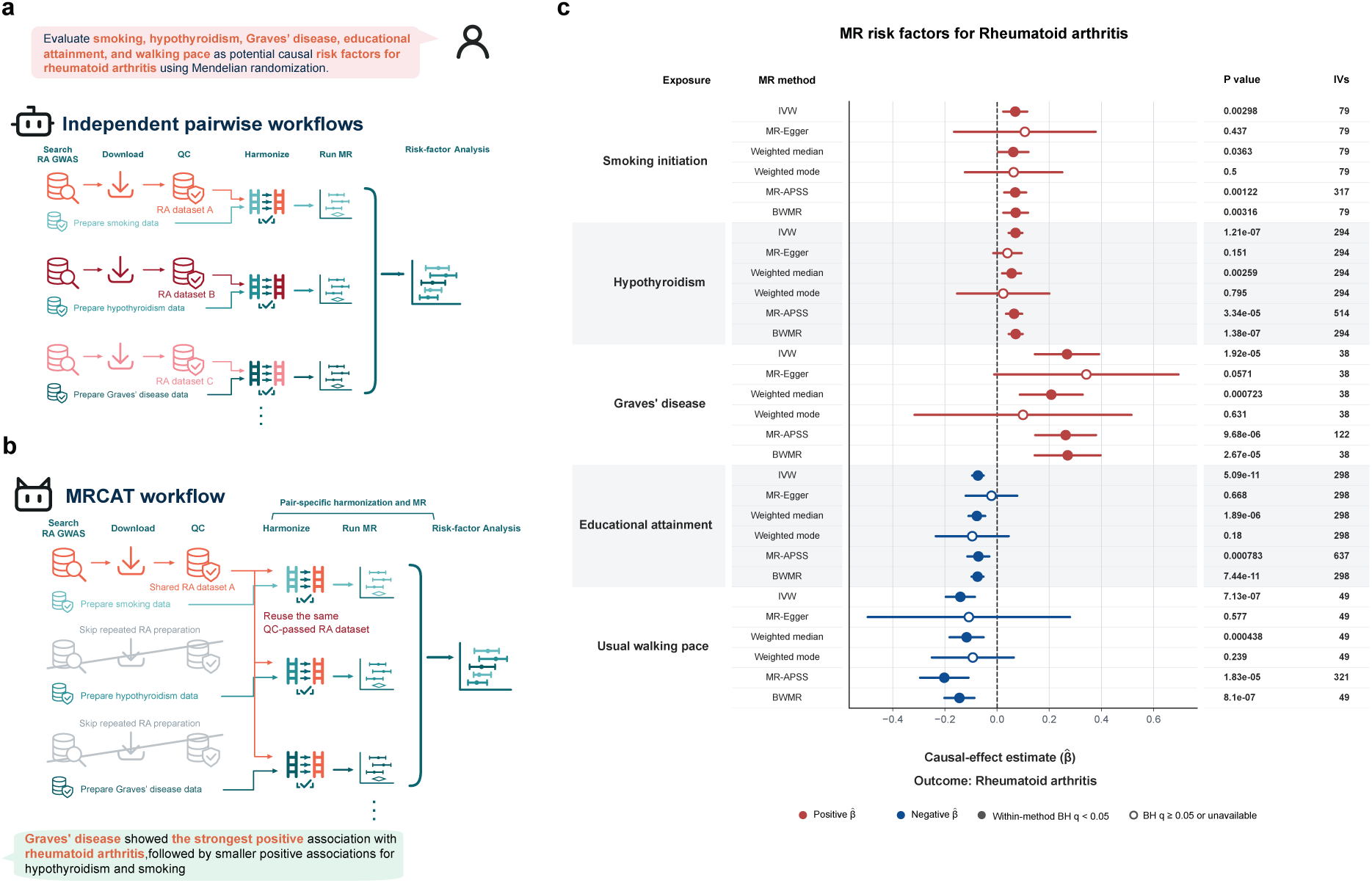
| Assessment of multiple candidate risk factors for rheumatoid arthritis with MRCAT. a, Schematic of independent pairwise workflows that repeat outcome-data acquisition and preparation, potentially selecting different rheumatoid arthritis (RA) datasets across exposures. **b,** MRCAT reuses a shared, quality-controlled RA dataset while retaining pair-specific harmonization and MR estimation, then combines results across exposures. **c,** Estimates for smoking initiation, hypothyroidism, Graves’ disease, educational attainment and usual walking pace in relation to RA, obtained using inverse-variance weighted (IVW), MR-Egger, weighted median, weighted mode, MR-APSS and Bayesian weighted MR (BWMR). Points denote effect estimates (β^^^) and horizontal bars denote 95% confidence intervals; the dashed line marks zero. Red and blue indicate positive and negative estimates, respectively. Filled points indicate Benjamini–Hochberg (BH) *q <* 0.05 and open points indicate *q ≥* 0.05. Columns report *P* values and the number of instrumental variables (IVs).

After completing the five exposure–outcome analyses, MRCAT consolidated 30 method-specific estimates— five exposures evaluated using six MR methods—into a unified risk-factor assessment (Supplementary Table 20). Each estimate remained linked to its source analysis and provenance. The resulting summary forest plot (Fig. 5c) was accompanied by a cross-method evidence matrix and pair-specific diagnostic plots, including funnel and leave-one-out plots (Supplementary Figs. 2–8). Thus, MRCAT coordinated analyses that shared suitable data-processing steps without treating their instruments, harmonization decisions or results as interchangeable.

The persistent workspace also supports follow-up analyses that change the causal direction. In the two-request scenario shown in Fig. 6a, the user first asks MRCAT to assess the effect of T2D on CHD and then requests an analysis of the reverse direction. Because the initial request specified CHD broadly, MRCAT selected a CAD GWAS after evaluating the phenotype and study metadata against the requested outcome. The Planner retained the completed forward analysis and assessed which quality-controlled datasets and analytical artifacts remained suitable for reuse. For the reverse analysis, it assigned CHD as the exposure and T2D as the outcome, independently selected CHD instruments and performed the harmonization and estimation steps required for this direction. The forward analysis could therefore provide reusable study-level data and a reference for comparison, but its instruments, harmonized inputs and estimates were not automatically transferred to the reverse analysis.

**Figure 6.**
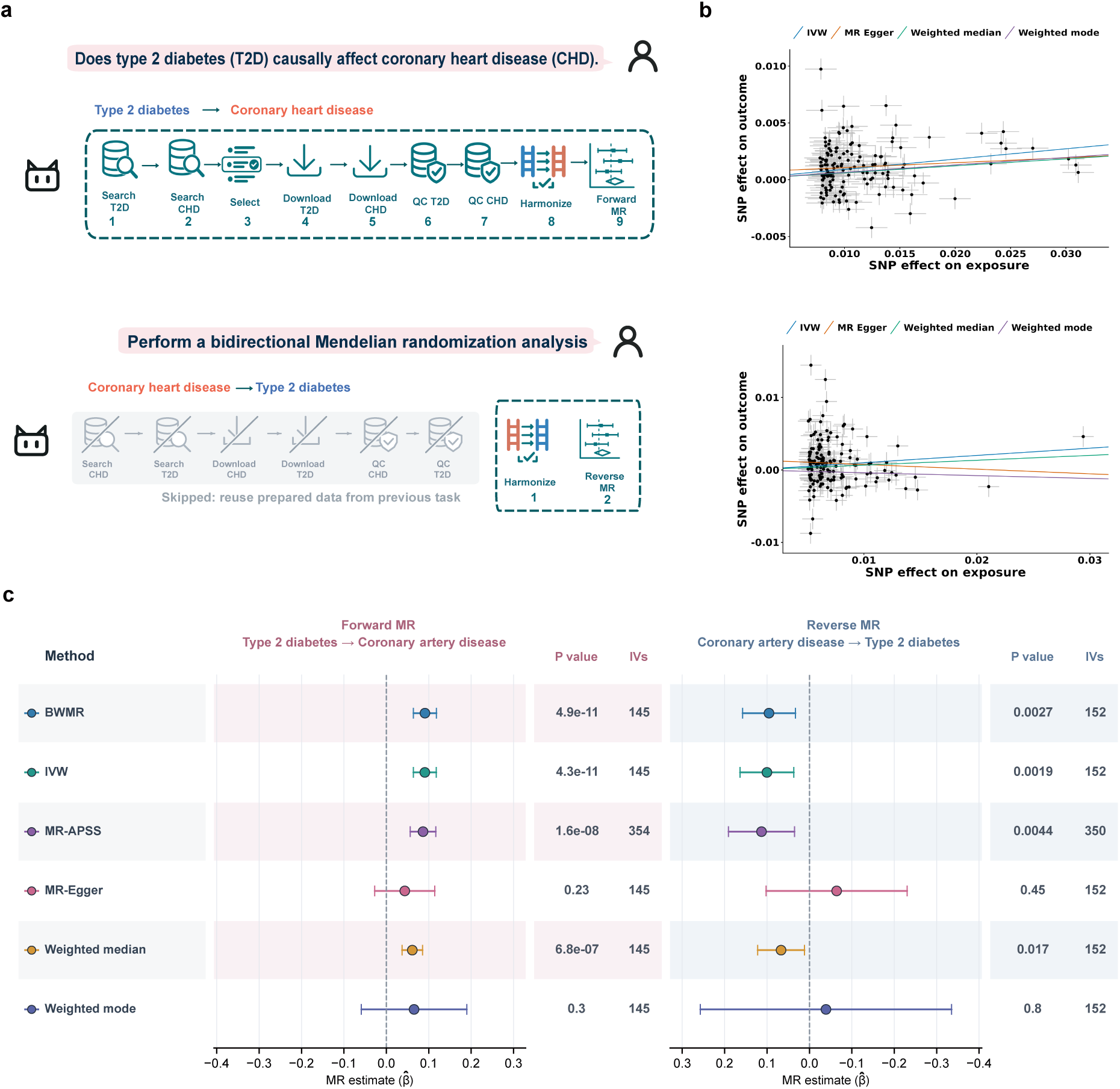
| Bidirectional MR using data retained from an earlier analysis. a, Schematic of the two-request workflow. The user first requests type 2 diabetes (T2D) to coronary heart disease (CHD) analysis, then requests bidirectional MR. In the numerical source session, CHD was represented by a coronary artery disease (CAD) GWAS. Grey crossed-out stages indicate data acquisition and quality control reused from the forward analysis; direction-specific input preparation is performed for the reverse analysis. **b,** MR scatter plots for the forward (T2D to CAD; top) and reverse (CAD to T2D; bottom) analyses; lines show IVW, MR-Egger, weighted median and weighted mode fits. **c,** Forward (T2D to CAD) and reverse (CAD to T2D) estimates from six MR methods. Points denote MR effect estimates (β^^^) and horizontal bars denote 95% confidence intervals. Dashed lines mark zero; the two directions use the same horizontal scale, and the reverse axis is mirrored. Columns give *P* values and IV counts.

MRCAT then applied the same six MR methods to the direction-specific inputs and combined the new estimates with the retained forward results to produce a bidirectional assessment (Supplementary Table 21; Supplementary Figs. 9–10). MR scatter plots displayed the SNP–exposure and SNP–outcome associations and fitted relationships separately for the forward and reverse analyses (Fig. 6b). A mirrored forest plot presented the estimates from both directions together with their confidence intervals and instrument counts (Fig. 6c). This workflow preserved the continuity of the investigation while ensuring that the change in exposure–outcome roles triggered the instrument selection, harmonization and estimation steps required for the reverse direction.

In summary, these examples demonstrate how MRCAT extends an ongoing MR investigation while maintaining methodological control. Its persistent workspace preserves research context and analytical provenance, whereas the Planner selectively reuses suitable outputs and identifies the pair-specific or directionspecific operations that must be repeated. MRCAT therefore supports integrated downstream assessments without assuming that previously generated data, instruments or estimates are automatically valid for a new exposure–outcome pair or causal direction.

## Discussion

Reliable automation of Mendelian randomization (MR) requires more than the ability to retrieve data, generate code and obtain an effect estimate. It also requires the agent to preserve methodological dependencies, identify when analytical prerequisites are unmet, and interpret results in a manner consistent with the evidence and MR assumptions. In our benchmark, comparator AI agents sometimes produced numerical estimates while omitting essential procedures or making causal claims that were not supported by the underlying analyses. We therefore developed MRCAT, a multi-agent framework that embeds MR-specific knowledge throughout the MR research cycle, from research-question formulation and data retrieval to analysis, review, interpretation and reporting.

MRCAT addresses procedural reliability through three coordinated mechanisms. Dependency-aware planning organizes analyses around their study-specific and method-specific prerequisites. Skill-guided execution combines established implementations with explicit decision rules, quality checks and stopping conditions. Expert-informed review evaluates failed executions, intermediate outputs and custom analyses and determines whether a step should be corrected, replanned or terminated. These mechanisms are particularly important for MR because errors in data matching, instrument selection, LD clumping or allele harmonization can propagate to downstream estimates and interpretations. MRCAT also distinguishes computational failure from a scientifically informative boundary, such as insuffiicient instruments, missing sample-size information or the absence of a suitable dataset. Thus, the system does not treat the production of an estimate as suffiicient evidence that an analysis has been completed appropriately.

MRCAT addresses inferential and reporting reliability by linking interpretation to the analytical record. Traceable evidence retrieval assembles estimates, diagnostics, figures, upstream outputs and provenance, while the Writer-Judge workflow checks reporting completeness, consistency between numerical results and their interpretation, and the support for causal claims. This design extends methodological scrutiny beyond execution to scientific communication. In particular, it helps prevent isolated nominal significance, agreement across methods or consistent effect directions from being presented as suffiicient evidence of a causal effect when the underlying procedures, uncertainty or replication evidence do not support such a conclusion. We evaluated these capabilities at several complementary levels. In the 30-query benchmark, MRCAT passed all nine prespecified procedural gates across the 26 executable queries and appropriately handled all four boundary cases. The comparator systems generated estimates for some queries without satisfying all procedural requirements, demonstrating that execution completion and scientific completion are distinct outcomes. Gate-level failures, including omitted or inappropriate LD clumping, incorrect allele handling and inconsistencies between executed and reported results, showed how domain-specific safeguards contributed to MRCAT’s performance. MRCAT also used fewer LLM calls and tokens than the comparator agents in matched scientifically completed runs, although these resource comparisons were conditional on successful runs and did not include the cumulative cost of earlier unsuccessful attempts.

Comparisons with expert-curated analyses provided a second level of validation. In the BMI-T2D MR-APSS analysis, MRCAT reproduced the expert-curated workflow and obtained numerical results consistent with the expert-curated analysis. By contrast, deviations in data preparation, background-parameter estimation, allele handling or LD clumping in comparator systems were accompanied by discrepant estimates or failed analyses. Across six validated negative-control tasks, MRCAT produced interpretations consistent with the expert-curated conclusion that the analyses did not support causal effects, whereas comparator agents sometimes interpreted nominal significance or agreement across methods as evidence of causality. These results support the value of combining procedural controls with evidence-grounded interpretation. They also illustrate why apparently consistent results across several MR methods do not necessarily provide independent confirmation when the methods share an upstream processing error.

The reporting case study further demonstrated how MRCAT operates across a longitudinal investigation rather than an isolated analysis. In the hexadecanedioate-CAD example, MRCAT integrated the original analysis, a FinnGen replication, diagnostic results and relevant literature into a traceable report. The report documented data sources, processing decisions, instrument selection, estimates and uncertainty, while distinguishing consistent effect directions from reproducible evidence for a protective causal effect. It also incorporated heterogeneity, influential-instrument analyses, the relaxed instrument-selection threshold and differences between the CAD outcome definitions. On this basis, the report characterized the findings as exploratory and concluded that the available evidence was insuffiicient to establish a protective causal effect. This example illustrates how report-level reliability depends not only on including the relevant results, but also on preserving their limitations and calibrating conclusions to the strength and reproducibility of the evidence.

MRCAT also supports downstream investigations through persistent workspaces and selective reuse. In the multi-exposure RA analysis, it reused suitable quality-controlled outcome data while independently selecting instruments and preparing each exposure-outcome pair. In the bidirectional T2D-CHD analysis, it retained the forward analysis for comparison and reused compatible study-level outputs, but independently selected instruments and performed the preparation required after reversing the analytical roles. These examples show that reuse can reduce redundant processing without treating previous instruments, harmonized inputs or estimates as automatically valid for a new method, exposure-outcome pair or causal direction. Such selective reuse may be particularly valuable for iterative MR investigations in which researchers progressively refine the research question or compare related analyses.

Several limitations should be considered. First, MRCAT currently focuses on MR workflows and does not replace expert judgment or establish that an inferred relationship is biologically causal. Its conclusions remain conditional on the quality of the source GWAS data [36], the validity of the encoded methods and the assumptions of the selected MR analyses [37, 38]. MRCAT cannot correct systematic problems in the underlying studies, and novel methods or analytical designs that are not represented in its Skill library may require custom code or human intervention. Second, the benchmark and expert-curated analyses covered a defined set of MR tasks and therefore cannot establish performance across all MR designs, populations, data resources or failure modes. Future evaluations should include a broader range of settings, such as multivariable MR, cis-MR, colocalization and complex ancestry structures. Third, the use of an LLM judge introduces a potential source of evaluation dependence, even though assessments were grounded in execution traces, intermediate files and prespecified criteria. Independent expert adjudication and inter-rater agreement should be incorporated into future evaluations. Finally, the current workspace supports analytical continuity but does not yet integrate complementary evidence streams such as fine-mapping, colocalization, functional genomics or experimental validation.

These limitations also define important directions for future development. Integrating complementary genetic, molecular and functional analyses could help researchers assess the biological relevance of MR findings and select appropriate follow-up studies, provided that the assumptions and limitations of each evidence source are represented explicitly [39]. User interaction could also be strengthened so that researchers can inspect intermediate decisions, revise analytical plans, challenge interpretations and determine when additional evidence is needed. Rather than replacing researchers, such interactions would support informed human oversight throughout the investigation [40].

Overall, MRCAT demonstrates how MR-specific expertise can be represented as a workflow that is dependencyaware, executable, verifiable, recoverable and traceable. Its results suggest that scientific agents should not be evaluated solely by whether they produce code or numerical outputs. They should also be assessed by whether they preserve analytical prerequisites, recognize scientifically meaningful limitations, reproduce verifiable results, communicate uncertainty and support evidence-grounded researcher judgment. For MR and other assumption-dependent biomedical analyses, this reliability-oriented approach may provide a foundation for more reproducible and transparent AI-assisted scientific research.

## Methods

### MRCAT system architecture

MRCAT is organized around a central Manager that coordinates three specialized agents: Search, MR and Report. The Manager interprets the request in the context of the ongoing investigation and routes it to the appropriate agent. Search agent retrieves and inspects external literature and data resources relevant to the research question. MR agent translates analytical requests into dependency-aware plans so that each step can obtain the required data and results from preceding steps; then performs the planned analyses through skill-guided execution, using MR methodological instructions and executable checks embedded in each Skill. To further support procedural reliability, expert-informed review handles execution problems and guides correction, replanning or termination when needed. Report agent retrieves relevant evidence from saved analytical results and integrates it with supporting literature through a Writer–Judge workflow to produce a traceable scientific report. This architecture enables end-to-end MR investigations, from information retrieval to analysis and reporting, while embedding safeguards for procedural reliability and inferential and reporting reliability. Furthermore, MRCAT complements session state with a persistent workspace that retains input data and analytical outputs. This facilitates reuse of compatible data and results in follow-up analyses, helping avoid unintended discrepancies that could arise from restarting the workflow and undermine subsequent conclusions.

#### Search agent

The Search agent retrieves the literature, GWAS data and related resources needed to start or extend an MR investigation. It helps users examine existing evidence, identify candidate datasets and inspect study information before analysis.

For literature queries, the agent formulates search queries and retrieves relevant sources. For GWAS discovery, it converts trait descriptions into search terms and aliases while preserving any supplied study identifiers and phenotype, source and population constraints. A CodeAct loop lets the agent call retrieval tools through code and inspect the returned records and source diagnostics. Missing metadata prompts inspection of the original study record, and an unsuccessful search prompts refinement with equivalent trait terminology. Retrieval failures are distinguished from a genuine absence of matching datasets, and findings are saved with source references.

We implemented three reusable GWAS Skills, available to both the Search and MR workflows. The gwas-search Skill queries OpenGWAS, the GWAS Catalog, Pan-UK Biobank and FinnGen (see Supplementary Methods and Results, Data sources and benchmark inputs) and returns candidates with study identifiers, phenotype descriptions, sample information and download metadata. The gwas-select Skill evaluates candidates against phenotype, source and population requirements and returns the selected records with its rationale. The gwas-download Skill retrieves the selected summary statistics together with study metadata and provenance.

### MR agent

#### Dependency-aware planning

When the Manager routes a new analysis request to the MR agent, the Planner generates a plan before execution begins. It uses the user’s request, Skill descriptions and input–output metadata, and available results to determine the required operations and their order. It distinguishes inputs already supplied by the user or available from compatible previous analyses from those that must be generated by preceding operations. To account for data dependencies, the Planner places operations that produce required inputs before those that consume them, guided by Skill metadata and MR-specific planning rules. These rules guide Skill selection and preparation according to method-specific requirements. For example, MR-APSS requires matching background parameters, while reverse-direction analyses require new instrument selection and pair preparation despite potentially reusable study-level quality-control outputs. The Planner assesses historical results against the requested studies, direction, population and method requirements before selecting them for reuse.

Each planned step records its identifier, analytical objective, selected Skill or custom-code designation, and references to historical results chosen for reuse. Before the plan is accepted, the runtime checks its structure, verifies that the selected Skills exist and confirms that historical reuse references point to saved results from completed steps.

#### Skill-guided execution

After the Plan is generated and confirmed, the MR subgraph enters the Executor, which processes the Plan items sequentially. For Skill-based items, the Executor generates the tool call, while runtime checks determine whether the item advances or enters Review. For custom items, the Executor iteratively writes, runs and revises code before submitting the analysis for Review.

When a Skill is assigned to the current item, the Executor receives the item description, user requirements, saved results from previous items and the complete SKILL.md guide. The guide explains the required inputs, parameter settings, analysis procedure, expected outputs, quality checks and boundaries. Following these instructions, the Executor generates code that calls the corresponding tool, which performs the analysis and saves structured results and output files. Methodological instructions guide context-dependent decisions, while executable checks enforce directly testable requirements, such as required input fields and minimum instrument counts. The workflow advances after checking that the expected tool has returned a successful saved result. A successful process exit alone does not establish completion: the saved result must originate from the selected tool and report a successful status. Failed execution, blocked status or invalid outputs trigger Review.

When no Skill is assigned to the current item, the Executor uses the Plan description as guidance, along with upstream results to write custom Python, R or Bash code. Execution outputs and errors are returned to the Executor, which uses this feedback to revise or extend the code until it submits the item for completion or reports an obstacle. Reusable custom outputs must be registered with references to the execution that produced them. Review then assesses the custom analysis against the user’s request, methodological requirements and saved execution evidence before allowing the workflow to continue. This design provides standardized execution for established MR Skills while retaining controlled flexibility for operations not covered by a compatible Skill.

#### Expert-informed review

Expert-informed review examines failed and blocked Skill executions, completed custom analyses and unresolved obstacles submitted by the custom Executor. Its inputs comprise the user’s request and constraints, the current plan item, execution evidence, and applicable methodological requirements. For custom analyses, Review assesses scientific scope, assumptions and supporting evidence, while technical debugging remains the Executor’s responsibility.

Review returns one of three decisions with a supporting reason: execute, replan, or finalize. Execute retains the current plan, returning incomplete work for correction and advancing past accepted custom analyses. Replan requests changes to analytical operations and their dependencies, identifying invalid outputs while preserving compatible completed work. Finalize initiates delivery when a supported boundary prevents further autonomous progress, summarizing available results, unresolved limitations and the requirements for continuation.

Domain expertise is encoded in the Review instructions as explicit rules for diagnosing MR-specific problems, selecting appropriate repairs and recognizing scientific boundaries such as unauthorized phenotype substitution or unmet prerequisites for a method. These rules preserve user-specified phenotypes, studies and methods while requiring corrections to address the underlying problem. For example, missing sample-size information must be resolved using study evidence rather than by relaxing the instrument-selection threshold. Repeated technical failures alone do not establish that suitable data are unavailable. Review thus distinguishes correctable execution failures from limitations that prevent scientifically justified analysis.

#### Shared MR Skill library

The shared Skill library comprises 26 reusable Skills: 3 for data acquisition, 2 for data preparation, 14 for MR estimation and 7 for downstream analysis. These cover study-level quality control, exposure–outcome pair preparation, causal estimation, sensitivity analyses and integration of completed results. Each Skill combines a Markdown guide with executable procedures, providing methodological requirements for planning, execution and review. The full catalog is provided in Supplementary Table 1.

For planning, Skill descriptions and input–output metadata summarize supported operations, required inputs and reusable outputs. The Planner uses this information to select compatible Skills, arrange dependent operations and identify existing results that can satisfy subsequent requirements. For execution, the complete guide specifies input requirements, parameter settings, calling instructions, expected outputs and methodological boundaries. Executable procedures perform the analysis and check directly testable requirements, while written guidance supports context-dependent decisions. For review, relevant sections of the Skills loaded during the task provide requirements for assessing inputs, diagnostics and methodological limitations. Review considers these requirements alongside saved execution evidence to identify unmet conditions and determine which outputs remain suitable for use. Dedicated review instructions guide the subsequent recovery decision. The library thus connects the requirements used to plan an analysis with its execution procedures and the criteria used to assess problems.

#### Report agent

The Report agent generates scientific reports from saved analytical results and relevant literature. It retrieves outputs from completed analysis steps, excludes missing, failed and invalidated results, and retains their task and execution identifiers. An LLM selects the analyses relevant to the reporting question from a catalog of saved steps and summaries. The system supplements this selection with associated GWAS quality-control results and available upstream evidence identified through recorded reuse references. Before drafting, the agent formulates background and result-comparison questions, retrieves PubMed records and assesses their abstracts for support and limitations. Analytical evidence, literature records and figures retain source identifiers, allowing reported findings to be traced to their underlying sources.

The Writer receives the selected evidence, assessed literature and figure catalog, together with MR-specific interpretation instructions and applicable STROBE-MR reporting guidance [25]. It produces a structured draft containing text, tables, figures and citations. The saved analytical record provides the basis for numerical results, while literature supports background discussion and comparison with published findings. Interpretation instructions require conclusions to account for effect scales, uncertainty, diagnostics and methodological assumptions. Programmatic checks validate the draft structure and ensure that figure and reference identifiers belong to the supplied catalogs.

The Judge compares the validated draft with the analytical and literature evidence, checking for substantive omissions, inconsistencies between results and interpretation, and unsupported causal claims. When revision is required, each issue specifies its location, the problem and the required correction. The Writer receives this feedback alongside the current draft and original evidence, then submits a revised draft for validation and renewed review. The workflow permits up to three revisions and ends early when the Judge accepts the report. A revision that neither obtains acceptance nor reduces the issue count is discarded, restoring the preceding draft and ending further revision. Reports are exported as Markdown and standalone HTML; drafts that have not passed review retain an explicit unaccepted status.

#### Persistent workspaces

MRCAT maintains structured state for each session, recording the current request, conversation, analysis plans, completed steps, task history, pending user input and output references. A persistent workspace stores input data and generated outputs, with executed code and execution logs saved alongside it. To retain useful context across turns, MRCAT summarizes user messages and execution observations into working memory organized by agent role and analysis task. For MR tasks, these summaries describe the task, confirmed facts, pending questions and recent progress. They are stored in session state and can be exported to memory.md, while the original transcripts and files remain available for inspection.

Within the same session, MRCAT interprets each follow-up request using saved state, recent messages and working summaries. It can continue unfinished work, revise a plan or create a new task using previous results. The Planner is instructed to assess whether saved results match the requested studies, analysis direction, population and method requirements. Selected results are referenced explicitly; runtime checks verify that references identify completed outputs and reject failed or invalidated results. For example, after an initial MR analysis, a user can request BWMR using previously prepared data that meet its input requirements, then request a report incorporating both analyses. Such reuse helps avoid unintended discrepancies in data and results that could arise from restarting the workflow and undermine subsequent conclusions.

### Benchmark protocols

Two complementary evaluations assessed workflow execution and boundary handling across diverse MR tasks, and concordance of numerical results and conclusions with expert-curated analyses.

#### 30-query benchmark (Fig. 2)

The benchmark comprised 30 queries, including 26 executable queries and four boundary queries. Executable queries requested MR analyses that could be completed under the specified conditions, whereas boundary queries introduced constraints requiring recognition of an analytical limitation and an appropriate response. The queries covered online data retrieval, analysis of local summary statistics, and combinations of local and online data, together with different requirements for MR methods. The complete queries and their classifications are provided in Supplementary Table 2.

*System comparison.* MRCAT, Claude Code (v2.1.220), Biomni (v0.0.8, A1 agent) and BioMedAgent (based on commit 74fa3f0) were evaluated on the 30-query benchmark using DeepSeek V4 Pro. The accompanying data archive records the exact request, input records for every system–query pair. Each system was provided with the same software packages. The tools supplied to BioMedAgent are listed in Supplementary Table 4. Software and package versions are listed in Supplementary Table 5.

*Evaluation metrics.* We used Codex (runtime version 0.164.0) with GPT-6.1 Sol to evaluate each system’s execution traces, intermediate result files and final reports. Using the same rubric for all systems (Supplementary Table 6), Codex assigned binary scores to G1–G9 for executable queries and B1 for boundary queries. Execution completion required at least one executed MR analysis yielding a numerical effect estimate. Reported estimates were checked against the corresponding analytical outputs. Query-level execution outcomes are shown in Supplementary Fig. 1a.

Boundary honesty was assessed using criterion B1, which could be satisfied through either of two alternative responses. A supported stop was credited when the limitation was identified, the affected analysis was paused, and the information or resources needed to continue were explained. Alternatively, a scientifically appropriate adaptation was credited when it was executed and its action, consequences and result scope were disclosed in the final solution report. Reported limitations and adaptations were checked against execution evidence. B1 outcomes were summarized separately across the four boundary queries and were not counted as scientific completion.

Procedural compliance was assessed using nine prespecified gates (G1–G9), each scored as 1 for pass or 0 for fail. The gates covered data matching, instrument selection, LD clumping, allele alignment, effectscale handling, method execution and reporting consistency. For reporting consistency, the methods, status and numerical results in the final solution reports were compared with the corresponding execution outputs. Requested analyses and additional analyses presented as evidence for the answer were assessed under the same rubric for all systems. The full rubric is provided in Supplementary Table 6, and query-level G1–G9 scores and B1 assessments are shown in Supplementary Fig. 1.

For each executable query, the procedure score was calculated as the sum of the nine gate scores. Scientific completion was defined more stringently, requiring all nine gates to be passed:

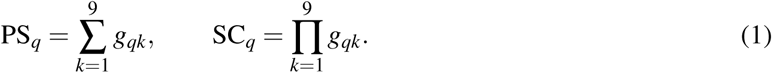

Here, *g_qk_ ∈ {*0, 1*}* indicates whether query *q* passed gate *k*. The mean procedure score was calculated across all 26 executable queries, including incomplete analyses, on a scale of 0–9. Scientific completion was summarized as the number of executable queries for which SC*_q_* = 1, out of 26.

*Resource use.* Resource use was measured for each system–query run by the total number of tokens processed and the number of LLM calls recorded during execution. MRCAT was compared separately with each comparator on matched executable queries. A query was included only when both systems passed all nine gates and had complete usage records. The same queries were used for both systems within each comparison, although the matched query sets differed between comparators. Usage records were matched to the sessions used for scoring. Reported costs covered the matched runs. Per-run measurements and token-accounting conventions are provided in Supplementary Table 7.

#### Comparisons with expert-curated analyses

Comparisons with expert-curated analyses (Fig. 3) used local GWAS summary-statistics datasets for one stepwise MR-APSS case and six negative-control exposure–outcome pairs (Supplementary Table 8). We also evaluated two additional positive-control pairs, high blood pressure–angina and LDL cholesterol–coronary artery disease (Supplementary Table 10). Trait labels were anonymized across all datasets to assess datadriven MR analysis while reducing reliance on prior knowledge of the named traits and their relationships. The complete queries are provided in Supplementary Table 9.

For the stepwise MR-APSS case, execution traces from all four systems were manually reviewed to compare the analytical steps and resulting outputs with the expert-curated workflow. For the six negative-control pairs, numerical results were extracted for each method reported in the final solution, and the stated conclusions were recorded as the system’s final interpretation. Numerical concordance was assessed against the corresponding method-specific expert-curated results, whereas interpretations were assessed against the negative-control expectations.

Expert-curated procedures and results for the stepwise case were obtained from the MR-APSS tutorial (https://raw.githubusercontent.com/YangLabHKUST/MR-APSS/master/MRAPSS_Rpackage_Tutorial. pdf). For the six negative-control pairs, method-specific expert-curated results were obtained from the publicly released MRbenchmarking result tables accompanying Hu et al. [31].

## Data availability

The GWAS data sources and access routes used in this study are described in Supplementary Methods and Results, “Data sources and benchmark inputs”, with dataset sources, study identifiers and download information provided in Supplementary Tables 8, 15 and 18. The complete queries for the 30-query benchmark and comparisons with expert-curated analyses are provided in Supplementary Tables 2 and 9, respectively. The MRCAT software package, example datasets and source code for reproducing the results are available at https://github.com/YangLabHKUST/MRCAT. MRCAT is available online at https://mrcat.science.

## Data Availability

The GWAS summary statistics used in this study were obtained from OpenGWAS (https://opengwas.io/), the NHGRI-EBI GWAS Catalog (https://www.ebi.ac.uk/gwas/), Pan-UK Biobank (https://pan.ukbb.broadinstitute.org/), and FinnGen (https://www.finngen.fi/en/access_results/), together with previously published datasets described in the Supplementary Information.

## Acknowledgements

This work was partly supported by the Innovation and Technology Commission (ITC-SKLNSD26SC01), Hong Kong Research Grants Council grants AoE/E-601/24-N, 16307322, 16302823, 16309424 and 16308925 and The Hong Kong University of Science and Technology start-up grants R9405 and Z0428 from the Big Data Institute.

## Author contributions

Y.J., F.Z., X.H., and C.Y. conceptualized and designed the study. Y.J. and F.Z. developed the methodology. Y.J. performed the analysis with the assistance of X. Hu and C.Y. X. Huang and Z.W. provided critical feedback during the study. Y.J., F.Z. and C.Y. wrote the manuscript.

